# Kidney Function Assessment Using Cystatin C and Serum Creatinine in Heart Transplantation Recipients: Implications for Valganciclovir Dosing

**DOI:** 10.1101/2023.11.22.23298924

**Authors:** Alberto Pinsino, Douglas L Jennings, Annamaria Ladanyi, Phuong Duong, Austin O Sweat, Ian Mahoney, Bruno Bohn, Ryan T Demmer, Koji Takeda, Gabriel T Sayer, Nir Uriel, Jay S Leb, Syed A Husain, Sumit Mohan, Paolo C Colombo, Melana Yuzefpolskaya

## Abstract

**Background:** Among heart transplantation (HT) recipients, the accuracy of serum creatinine (sCr)-based estimated glomerular filtration rate (eGFR) may be limited by fluctuations in muscle mass. Cystatin C (cysC) is less influenced by muscle mass; however, its levels may increase with obesity and steroid use. Herein, we: i) longitudinally compared changes in eGFRcysC and eGFRsCr among HT recipients; ii) investigated the association of body mass index (BMI), steroid use and muscle mass with discrepancies between eGFRs; and iii) explored the implications of eGFRcysC use on valganciclovir (VGC) dosing.

**Methods:** cysC and sCr were measured in 294 blood samples obtained from 80 HT recipients. Intra-individual differences between eGFRs (eGFRdiff_cysC-sCr_) were calculated. Negative eGFRdiff_cysC-sCr_ values correspond to eGFRsCr > eGFRcysC and positive values to eGFRcysC > eGFRsCr. In a patient subset (n=21), pectoralis muscle measures were derived from computed tomography scans.

**Results:** Marked differences between eGFRcysC and eGFRsCr were observed, particularly early post-HT (1-week post-HT, median eGFRdiff_cysC-sCr_ −28 ml/min/1.73 m^2^). eGFRcysC demonstrated stability following a transient post-operative decline, while eGFRsCr decreased in the first year post-HT. Lower BMI and higher prednisone dose displayed a modest association with more negative eGFRdiff_cysC-sCr_ values. Conversely, pectoralis muscle measures indicative of greater muscle mass and better tissue quality exhibited a stronger association with more positive eGFRdiff_cysC-sCr_ values. eGFRcysC use would have resulted in VGC dose adjustment in 46% of samples, predominantly towards lower doses.

**Conclusions:** Among HT recipients, eGFRcysC and eGFRsCr markedly differ with implications for VGC dosing. The observed discrepancies may reflect changes in body composition and steroid use.

## INTRODUCTION

Kidney dysfunction is a frequent complication following heart transplantation (HT) and is associated with increased morbidity and mortality^1–3^. Several factors are implicated in the pathogenesis of kidney dysfunction, including patients’ comorbidities (e.g., diabetes, hypertension) and HT-related factors (e.g., post-operative kidney injury, prolonged calcineurin inhibitor use)^4,5^. An accurate assessment of kidney function is essential both pre- and post-HT for identification of patients who may benefit from concomitant kidney transplantation^6^ and to guide medical therapy (e.g., renally-dosed medications, calcineurin inhibitor sparing regimens). However, serum creatinine (sCr)-based equations utilized to calculate estimated glomerular filtration rate (eGFR) may have limited accuracy among HT recipients^7^.

In advanced heart failure (HF), high rates of sarcopenia and malnutrition lead to decreased sCr generation, potentially resulting in overestimation of kidney function^8^. Following HT, the interpretation of changes in sCr levels is further complicated by several GFR-independent factors, such as post-operative immobility causing further muscle mass loss and changes in nutritional status.

Cystatin C (cysC) is an alternative eGFR marker which is less influenced by muscle mass, nutritional status, and demographics than sCr^9^. Cystatin C-based equations have shown superior accuracy in the general population^10,11^, and across various patient subgroups, including those with HF^12,13^. However, prior research investigating cysC-based assessment of kidney function in a HT population is limited to a single report in a small cohort of predominantly White males^14,15^. Notably, the potential impact of cysC confounders, such as changes in body mass index (BMI)^16^ and steroid use^17,18^, which are relevant in this patient population^19^, has not been previously explored. Furthermore, the implications of cysC-based estimates for post-HT medication dosing remain to be defined.

Valganciclovir (VGC), an antiviral medication commonly used for prophylaxis against cytomegalovirus (CMV) infection in HT, requires dose adjustments based on kidney function. Inappropriately high VGC dose in relation to kidney function may contribute to the high rate of leukopenia observed among HT recepients^20–22^. The management of this complication often requires dose reduction or discontinuation of vital medications, including immunosuppressants, potentially increasing morbidity and mortality^22^. Therefore, an accurate estimation of GFR is of paramount importance to prevent VGC toxicities and optimize outcomes.

In a cohort of HT recipients, we aimed to: i) longitudinally compare cysC- and sCr-based kidney function estimates at multiple time points; ii) investigate the potential contributions of BMI, steroid use and muscle mass to the discrepancies observed between cysC- and sCr-based estimates; and iii) explore the implications of cysC-based estimates for VGC dosing.

## MATERIALS AND METHODS

### Study design and data collection

We identified all patients who received HT at Columbia University Irving Medical Center between June 2016 and April 2019. The multi-organ and retransplant recipients were excluded. Cystatin C and sCr were measured simultaneously at the following time points: pre-HT, and serially post-HT: 1 week, 1 month, 2 to 3 months, 5 to 6 months, 1 year. Clinical data were obtained from electronic medical records. All participants provided informed consent, and the study was approved by the institutional review board.

### Estimated glomerular filtration rate

In the primary analysis, eGFR was calculated using the 2012 CysC-based (eGFRcysC) and the 2021 sCr-based (eGFRsCr) Chronic Kidney Disease Epidemiology Collaboration (CKD-EPI) equations^10,11^. Additional analyses included eGFR calculated with the 2021 CKD-EPI equation combining both markers (eGFRsCr+cysC)^11^. For each concurrent cysC and sCr measurement, the difference between the two estimates (eGFRdiff_cysC-sCr_) was calculated as the difference between log-transformed eGFRcysC and eGFRsCr, which is equivalent to the log-transformed ratio eGFRcysC/eGFRsCr^23^. Negative values of eGFRdiff_cysC-sCr_ correspond to eGFRsCr > eGFRcysC and positive values to eGFRcysC > eGFRsCr.

### Pectoralis muscle measures

Pectoralis muscle measures were performed in a patient subset with computed tomography (CT) scan obtained for clinical purposes within 14 days of cysC and sCr sampling. Methods were adapted from previous studies^24,25^. In brief, the analysis was performed using Aquarius iNtuition Viewer (TeraRecon Inc., San Mateo, CA) by a skilled radiologist blinded to cysC and sCr values. Unilateral cross-sectional pectoralis muscle area (cm^2^) and attenuation (PHUm_;_ pectoralis muscle mean Hounsfield units) were measured on the first axial slice above the superior aspect of the aortic arch. To account for body size, the pectoralis muscle index (PMI; cm^2^/m^2^) was obtained by dividing unilateral cross-sectional pectoralis muscle area by the height in square meters. Muscle attenuation (PHUm) represents a proxy for the tissue quality, where lower values indicate intramuscular fatty infiltration associated with impaired muscle function^26^.

### Valganciclovir dosing and leukopenia events

In accordance with the manufacturer’s recommendation, VGC dosing is based on creatinine clearance (CrCl) calculated with the Cockcroft-Gault equation^27^. Among patients receiving VGC at 1, 2 to 3 and 5 to 6 months post-HT, the implications of different kidney function estimates on VGC dosing were examined. To allow for comparison with CrCl, eGFRcysC, eGFRsCr and eGFRsCr+cysC values were re-expressed in mL/min by multiplying by body surface area derived from the DuBois formula divided by 1.73 m^2^ ^28,29^. In addition, the association of different GFR estimates with leukopenia events was explored. Leukopenia was defined as a white blood cell count <4 × 10^9^/l within 1-month of sampling and after excluding other identifiable causes (e.g., CMV infection, bacterial sepsis).

Only samples collected at 1, 2 to 3 and 5 to 6 months post-HT contributed to this analysis, due to the following: i) the rate of leukopenia in the first month post-HT is low^22^; and ii) our institutional protocol recommends discontinuing VGC after 6 months of prophylaxis.

### Statistical analysis

Kidney function estimates were log-transformed to ensure normality. Linear mixed models regressed cysC- and sCr-based estimates as well as eGFRdiff_cysC-sCr_ longitudinally at different time points. Patients were modelled as random effects to account for within-person correlation of repeated measurements. All models had an exchangeable correlation structure and included age and sex. To more rigorously control for time-invariant confounders, we performed sensitivity analyses restricted to patients who had samples available at both: pre-HT and 1-week post-HT; pre-HT and 1-month post-HT; 1-week and 5 to 6 months post-HT; and 1-month and 5 to 6 months post-HT. Comparisons between kidney function estimates were performed using the Wilcoxon signed-rank test. Using linear mixed models with the same correlation structure and covariates as described above, we analyzed: i) in the entire study population, longitudinal changes in BMI and prednisone dose, and their association with cysC- and sCr-based estimates as well as eGFRdiff_sCr-cysC_; ii) among patients with available CT scan, the association of PMI and PHUm with cysC- and sCr-based estimates as well as eGFRdiff_cysC-sCr_.

Generalized linear models with an exchangeable correlation structure were used to investigate the association of different kidney function estimates with leukopenia. Each observable period began at the time of sampling and continued up to the occurrence of leukopenia. Censoring took place at the time of: i) leukopenia attributable to an identifiable cause (e.g., CMV infection, bacterial sepsis); ii) any change in VGC dosing or VGC discontinuation; and iii) 1 month from sampling. All models were adjusted for white blood cell count at the time of sampling, VGC dose, mycophenolate dose, and sulfamethoxazole-trimethoprim use. A two-sided p-value <0.05 was considered statistically significant. All analyses were performed using SAS Version 9.4 (SAS Institute, Cary, NC).

## RESULTS

### Cystatin C-versus serum creatinine-based estimated glomerular filtration rate

Among 80 HT recipients, 294 blood samples were collected. Baseline characteristics are reported in **Table 1**.

We found significant discrepancies between eGFRcysC and eGFRsCr over the study follow-up (**Figure 1, Table S1**). Estimated GFRcysC markedly decreased from pre-HT to 1-week post-HT and subsequently increased with time post-HT. In contrast, eGFRsCr remained stable early post-HT, but showed a decline at 5 to 6 months that was sustained at 1-year post-HT (**Table S1**). The difference between the two estimates (eGFRdiff_cysC-sCr_) widened early after HT, with cysC yielding markedly lower estimates than sCr at 1-week and 1-month post-HT (median eGFRdiff_cysC-sCr_ −28 [interquartile range {IQR}: −40 to −18] and −22 [IQR: −31 to −6] ml/min/1.73 m^2^, respectively). This early discrepancy between eGFRcysC and eGFRsCr attenuated over time, with no statistically significant differences observed at 5 to 6 months and at 1-year post-HT.

**Figure 1.**
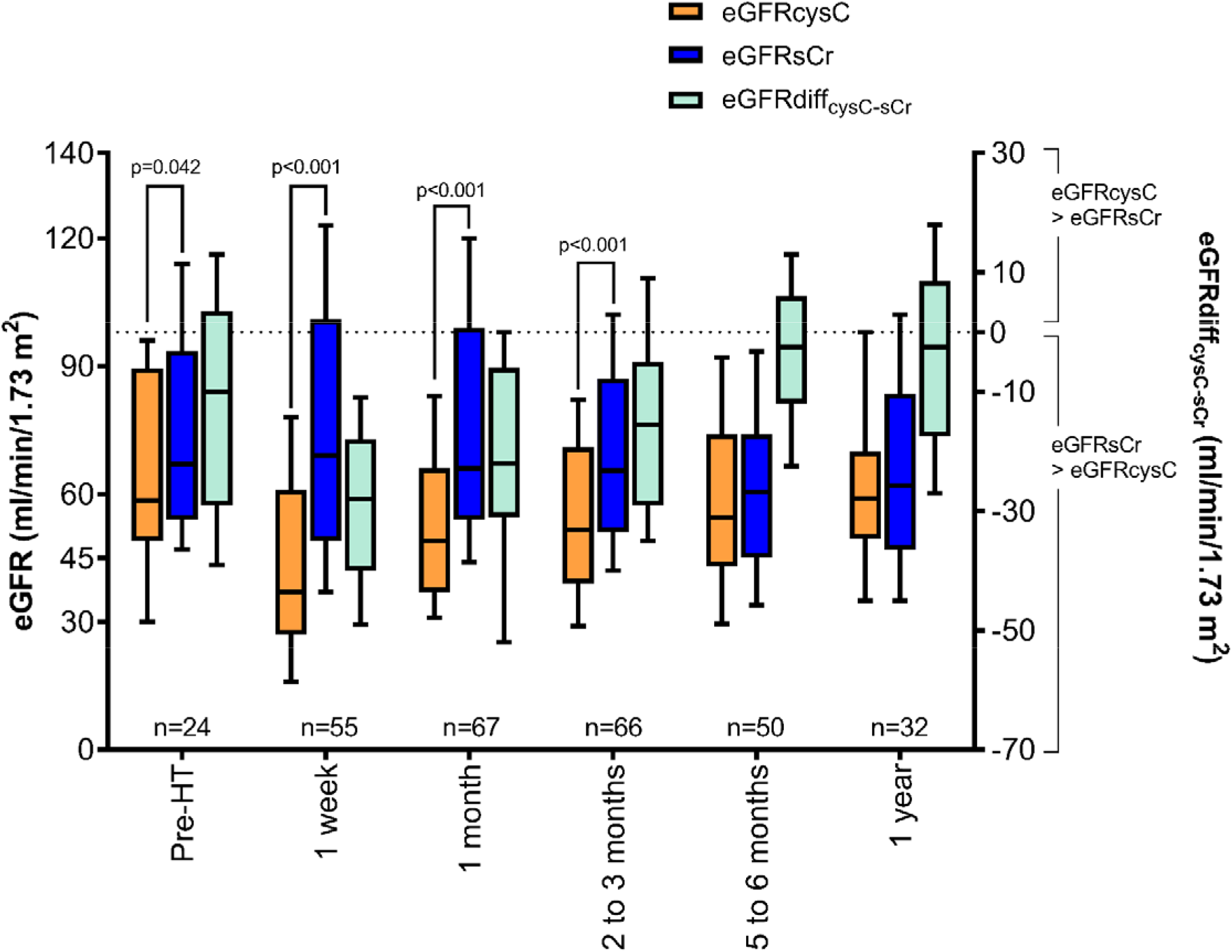
Kidney function estimates and the difference between cystatin C- and serum creatinine-based estimated glomerular filtration rate (eGFRdiff_cysC-sCr_) across time points. Boxes and whiskers represent the interquartile range and the 10th–90th percentile range, respectively. cysC indicates cystatin C; eGFR, estimated glomerular filtration rate; HT, heart transplant; sCr, serum creatinine.

**Table 1.**
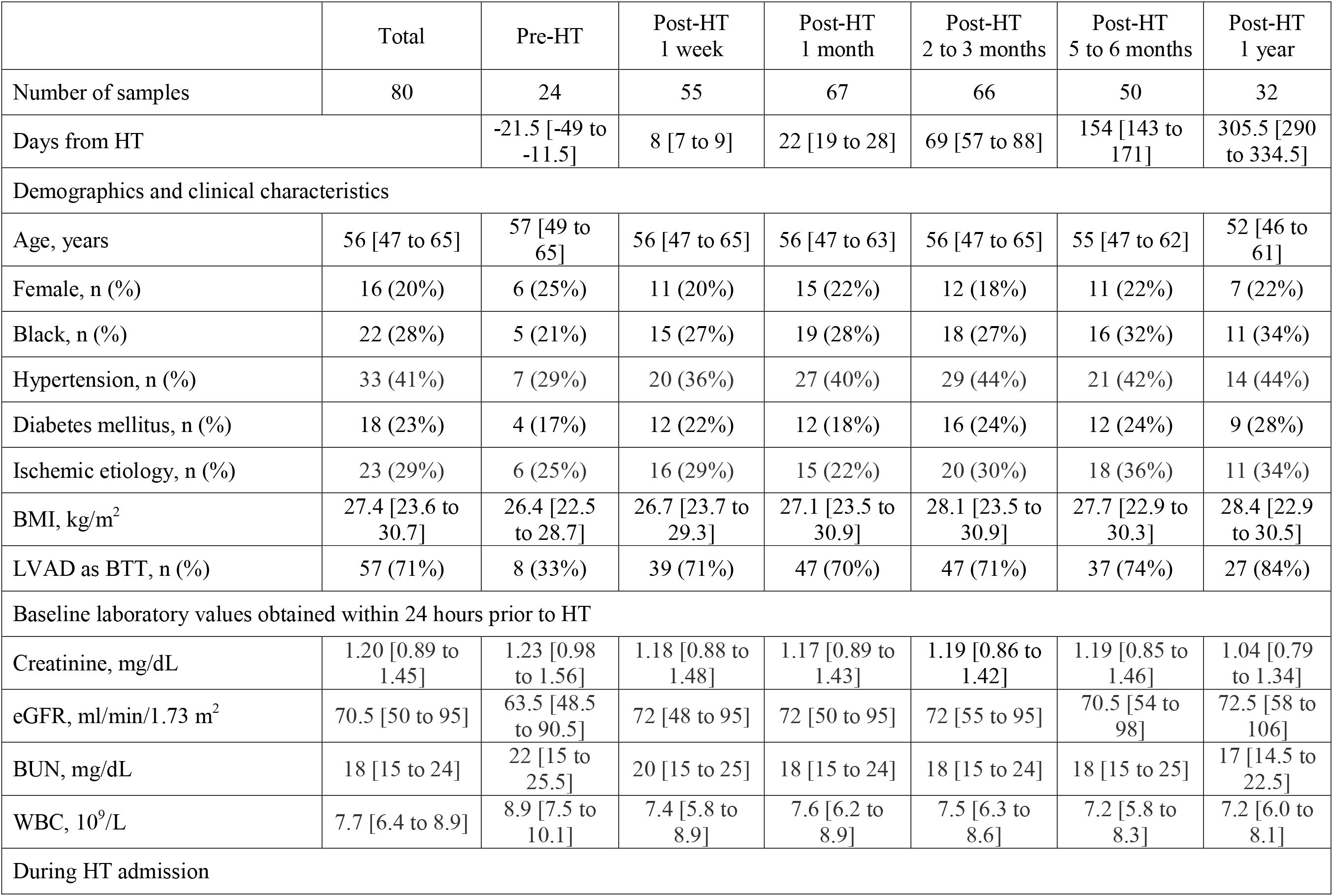
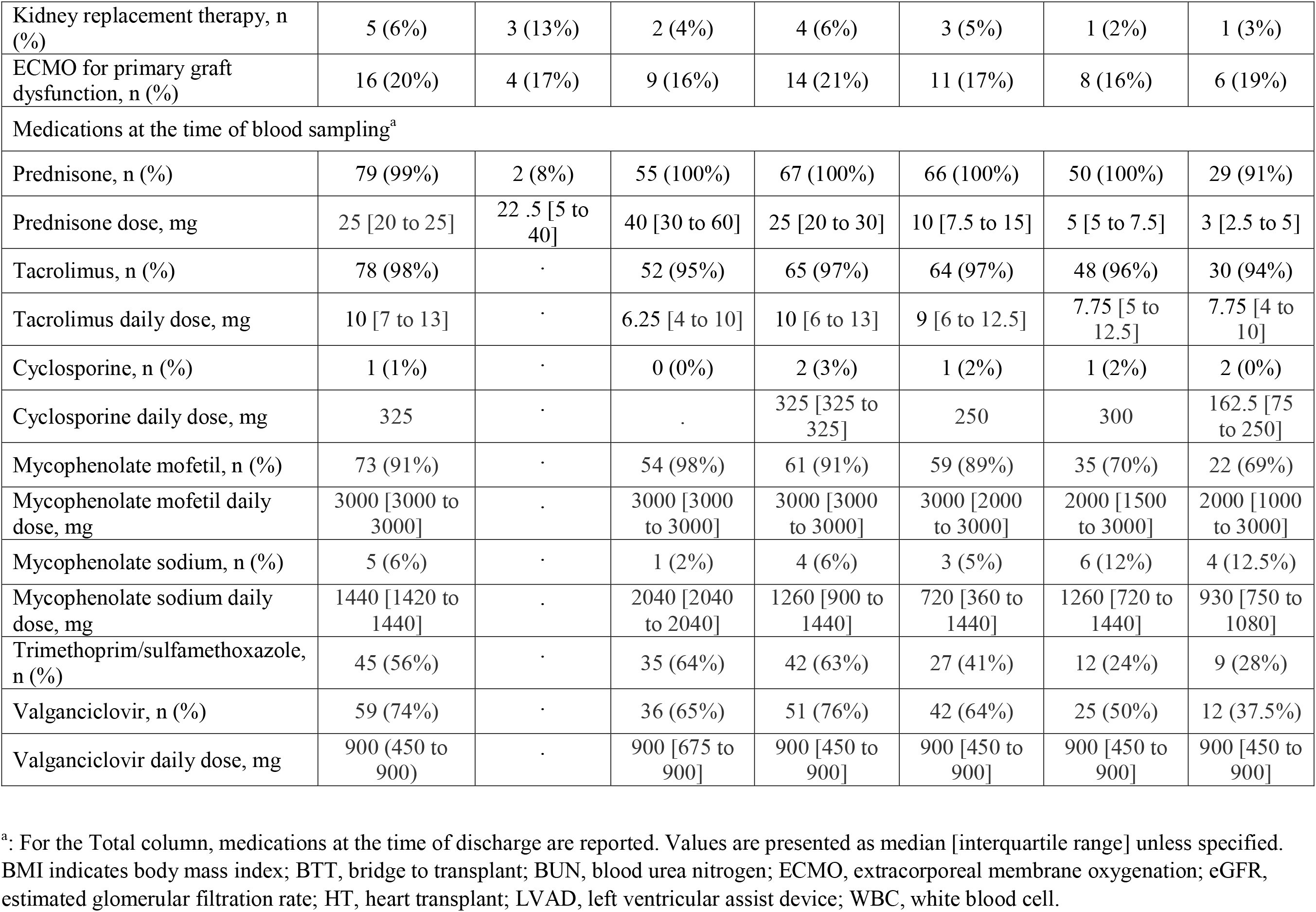
Baseline characteristics and drug regimens of study participants stratified by time of sampling.

Sensitivity analyses restricted to patients with paired samples were consistent with the main analysis (**Figure S1**). Estimated GFR calculated with an equation combining both glomerular filtration markers (eGFRsCr+cysC) yielded values which were intermediate between eGFRcysC and eGFRsCr (**Table S1, Figure S2**). Following a transient decline at 1-week, eGFRsCr+cysC remained stable post-HT.

Compared with eGFRsCr, eGFRcysC reclassified majority of samples to a different eGFR category, predominantly towards more advanced kidney dysfunction (**Figure 2**). Specifically, reclassification to worse eGFR category occurred in 10 (42%) patients pre-HT, 48 (87%) patients at 1-week post-HT and 48 (72%) patients at 1-months post-HT (**Table S2**). Overall, 38 (13%) samples exhibited an eGFR <30 mL/ min/1.73 m^2^ based on cysC compared with only 4 (1%) based on sCr (**Figure 2**). The reclassification resulting from eGFRsCr+cysC is shown in **Table S3**.

**Figure 2.**
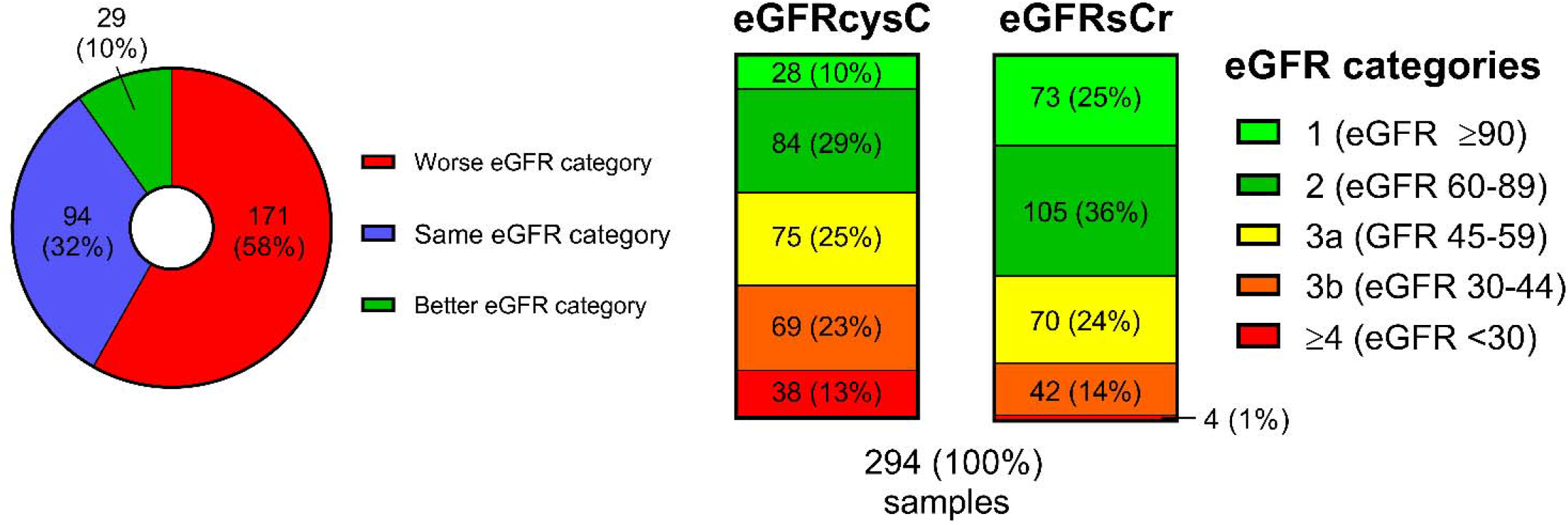
Reclassification of estimated glomerular filtration rate (eGFR) categories with cystatin C. Data are presented as number of samples and percentage of the total samples (%). cysC indicates cystatin C; sCr, serum creatinine.

### Body mass index, prednisone dose and pectoralis muscle measures: association with kidney function estimates and eGFRdiff_cysC-sCr_

Post-HT changes in BMI and prednisone dose are depicted in **Figure 3**. After an initial decline at 1 month, BMI increased and was significantly higher than pre-operative values at 5 to 6 months and 1-year post-HT. Prednisone was administered at a median dose of 40 mg/day at 1-week post-HT and subsequently tapered. Higher BMI was associated with lower eGFRsCr and eGFRsCr+cysC but had no significant association with eGFRcysC (**Table S4**). Prednisone dose was not associated with any cysC or sCr-based estimates.

**Figure 3.**
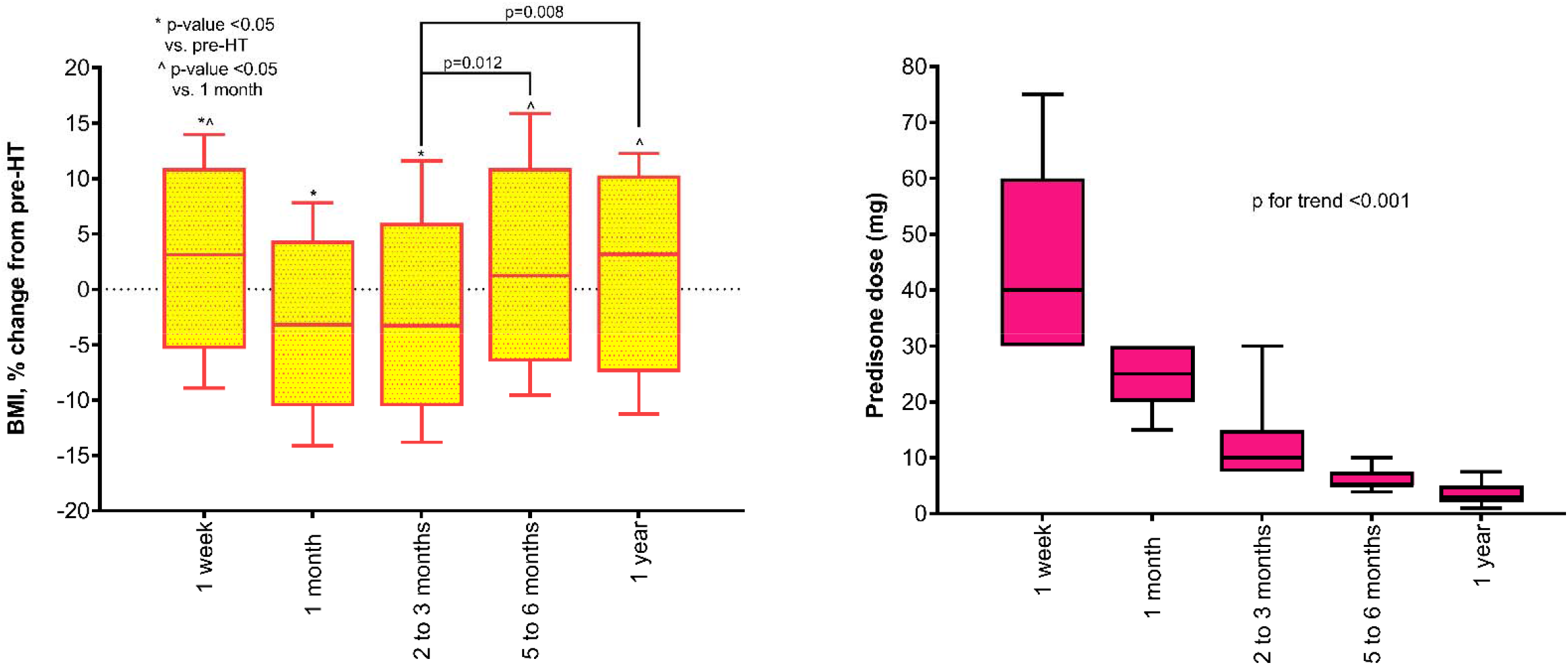
Longitudinal changes in body mass index and prednisone dose. Boxes and whiskers represent the interquartile range and the 10th–90th percentile range, respectively. P-values were obtained using linear mixed models adjusted for age and sex. BMI indicates body mass index; HT, heart transplant.

We found a modest but statistically significant association of BMI and prednisone dose with eGFRdiff_cysC-sCr._ A 10% increase in BMI was associated with a 2.1% change in eGFRdiff_cysC-sCr_ towards more positive values (95% confidence interval [CI]: 0.2 to 4.1%, p=0.029). For each 10% increase in prednisone dose, there was a 1.5% change in eGFRdiff_cysC-sCr_ towards more negative values (95%CI: 0.8 to 2.2%, p<0.001). Similar results were obtained in models including both BMI and prednisone dose (**Table S4**).

Baseline characteristics of the subset of study participants with available pectoralis muscle measures are shown in **Table S5.** Among 21 patients with a total of 40 CT scans obtained at a median of 3 (IQR: 1 to 8) days from sampling, average PMI was 4.9 ± 2.1 cm^2^/m^2^ and average PHUm was 27 ± 14 mean Hounsfield units. Higher PHUm was associated with higher eGFRcysC (β=0.1154, 95%CI: 0.0017 to 0.0214, p= 0.023). No other association between pectoralis muscle measures and individual cysC- or sCr-based eGFR reached statistical significance. Higher PMI and PHUm were associated with more positive eGFRdiff_cysC-sCr_ values (**Figure 4).** In adjusted models, for each 1-standard deviation (SD) increase in PMI and PHUm, eGFRdiff_cysC-sCr_ changed towards more positive values by 12.9% (95%CI: 1.7 to 25.2%, p=0.023) and 17.8% (95%CI: 6.6 to 30.2%, p=0.002), respectively. Further adjustment for BMI and prednisone dose did not significantly alter the association between pectoralis muscle measures and eGFRdiff_cysC-sCr_ (data not shown).

**Figure 4.**
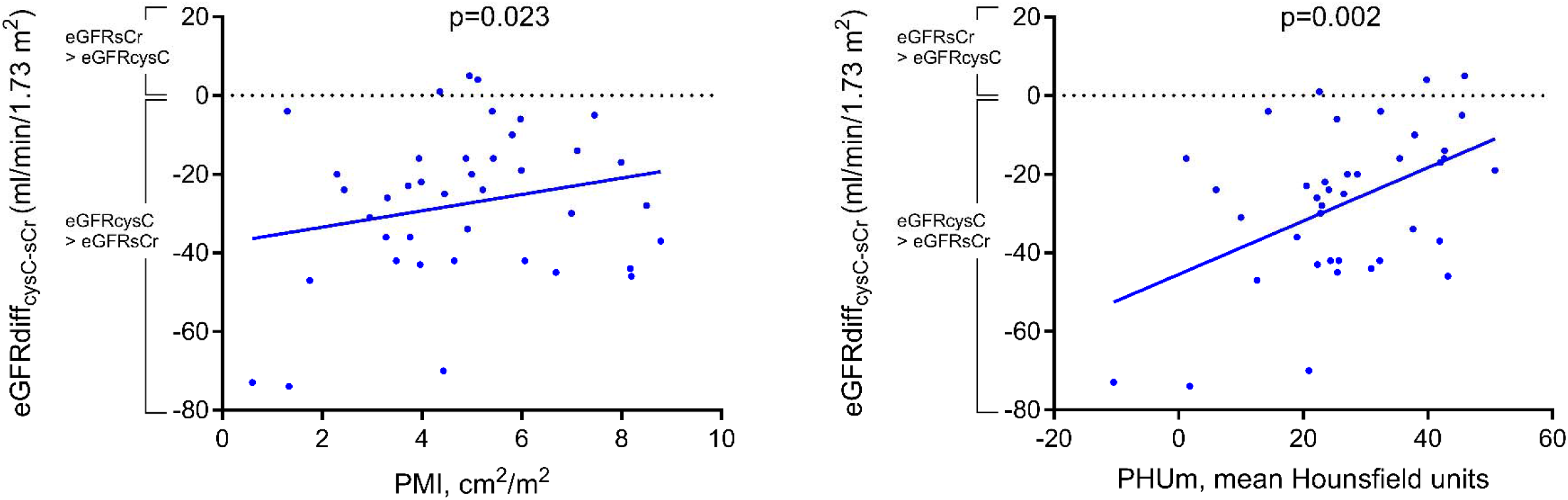
Association of pectoralis muscle measures with the difference between cystatin C- and serum creatinine-based estimated glomerular filtration rate (eGFRdiff_cysC-sCr_). P-values were obtained using linear mixed models adjusted for age and sex. cysC indicates cystatin C; eGFR indicates estimated glomerular filtration rate; PHUm, pectoralis muscle mean Hounsfield units; PMI, pectoralis muscle index; sCr, serum creatinine.

### Implications of different kidney function estimates for valganciclovir dosing and association with leukopenia

The baseline characteristics of the 57 study participants receiving VGC at 1, 2 to 3, and 5 to 6 months post-HT are presented in **Table S6**. Over 113 total samples, the use of eGFRsCr for VGC dosing would have resulted in dose adjustment in 30 (26%) samples; of those, 16 (14%) would have been *underdosed* and 14 (12%) *overdosed* (**Figure 5**).

**Figure 5.**
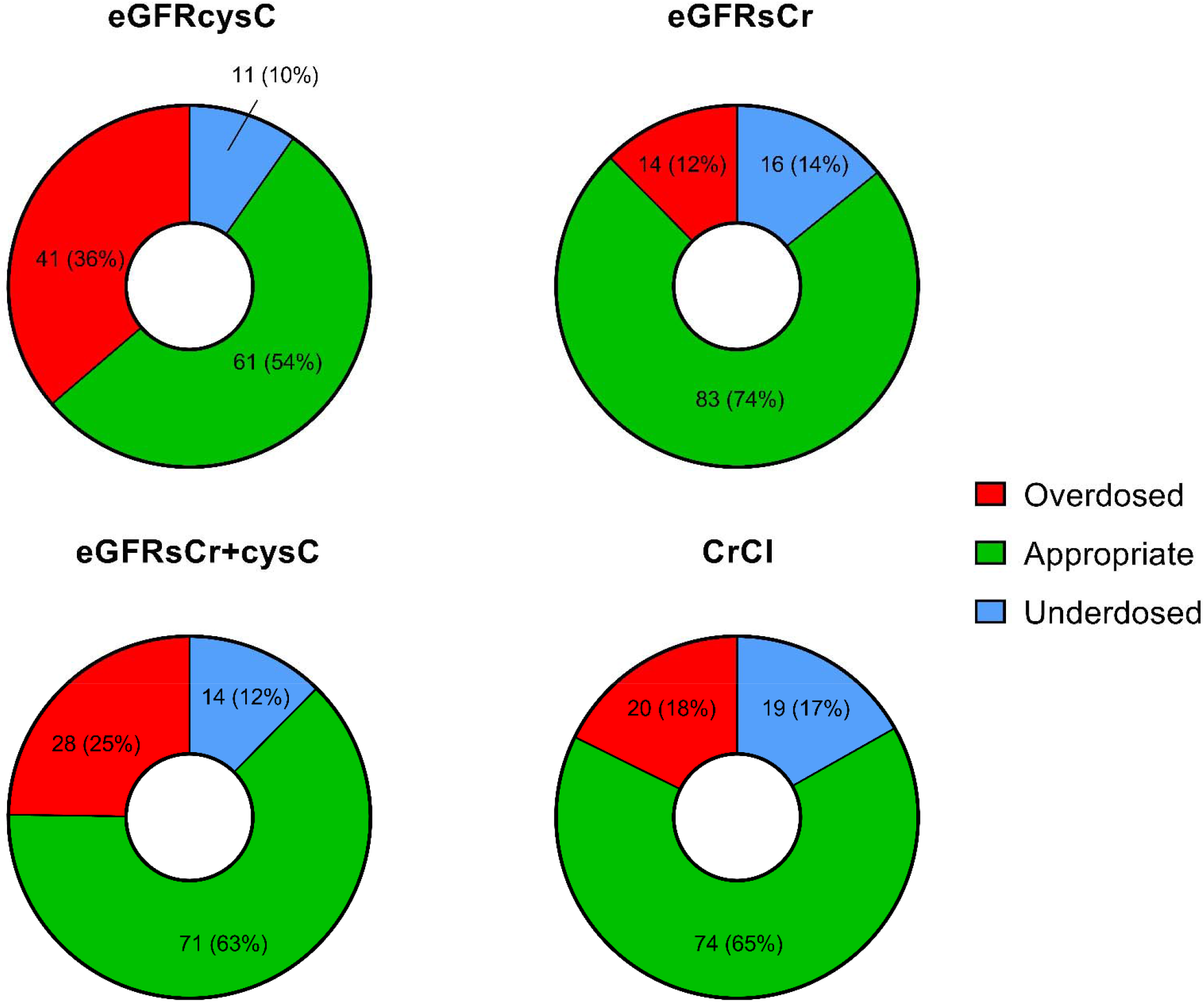
Dose adjustments based on estimates of kidney function compared to administered dose. CrCl indicates creatinine clearance; cysC, cystatin C; eGFR, estimated glomerular filtration rate; sCr, serum creatinine.

The use of eGFRcysC for VGC dosing would have resulted in dose adjustment in 52 (46%) samples; of those, 11 (10%) would have been *underdosed* and 41 (36%) *overdosed*. Of note, only 74 (65%) of samples had appropriate VGC dosing per the manufacturer’s recommended approach based on CrCl. Due to the marked discrepancies between cysC- and sCr-based estimates observed early post-HT, VGC dose adjustment resulting from eGFRcysC use would have been observed mainly within the first 3 months post-HT (**Figure S3**).

A total of 30 leukopenia events occurred among 27 individual patients. In adjusted models including VGC dosing, for each 1-SD decrease in eGFRcysC, eGFRsCr+cysC and CrCl, the risk for leukopenia approximately doubled (**Table 2**). The association of eGFRsCr with leukopenia did not reach statistical significance (p=0.083).

**Table 2.** Association of estimates of kidney function with leukopenia.

|  | Leukopenia<br>Samples n=113, events n=30 |  |
| --- | --- | --- |
|  | OR (95%CI) | p-value |
| eGFRcysC | 2.00 (1.15 to 3.47) | 0.014 |
| eGFRsCr | 1.75 (0.93 to 3.29) | 0.083 |
| eGFRsCr+cysC | 2.02 (1.07 to 3.82) | 0.03 |
| CrCl | 2.28 (1.05 to 4.99) | 0.038 |

Odds ratio per 1-standard deviation decrease in eGFR. All models were adjusted for white blood cell count at the time of sampling, valganciclovir dose, mycophenolate dose, and sulfamethoxazole-trimethoprim use. cysC indicates cystatin C; CrCl, creatinine clearance; eGFR, estimated glomerular filtration rate; OR, odds ratio; sCr, serum creatinine

## DISCUSSION

In a cohort of 80 HT recipients, we found marked discrepancies between cysC- and sCr-based estimates, particularly in the early post-HT period. While eGFRsCr remained stable in the post-operative period and declined afterwards, eGFRcysC worsened early post-HT but improved long-term. Body mass index and prednisone dose were modestly associated with the difference between the two estimates (eGFRdiff_cysC-sCr_). However, in a patient subset with available CT scan, a stronger association was present between pectoralis muscle measures indicative of greater muscle mass and better tissue quality and more positive eGFRdiff_cysC-sCr_ values. The use of cysC-based estimates would have led to frequent dose adjustments for VGC, primarily favoring dose reductions.

The measurement of clearance of exogenous markers (e.g., iohexol) represents the gold standard for kidney function assessment. However, its utility in clinical practice is limited by complexity of administration and cost, making estimates based on endogenous markers such as cysC and sCr more common. Despite the traditional reliance on sCr for kidney function assessment, two major nephrology societies have advocated for the routine use of cysC, citing its potential for greater accuracy and its independence from race^30^. Yancy et al. have similarly endorsed wider implementation of cysC-based estimates in the care of patients with HF^31^. In a recent consensus statement on heart-kidney transplantation, Kobashigawa et al. suggested the use of the CKD-EPI equation combining sCr and cysC^6^. Additionally, the most recent guidelines from the International Society for Heart and Lung Transplantation present cysC as an alternative for kidney function monitoring in HT recipients^32^. Our findings indicate that cysC- and sCr-based estimates differ significantly and cannot be used interchangeably.

A marked difference between cysC and sCr-based estimates occurs when factors unrelated to kidney function differentially affect the two markers. As previously reported by our group^23,33^, the discrepancy between eGFRs widens in the setting of worsening HF, potentially due to variations in sCr confounders such as muscle mass and nutritional status. The present study supports the hypothesis that muscle mass represents a major determinant of eGFRdiff_cysC-sCr_ among HT recipients. This is in line with prior evidence in patients implanted with left ventricular assist device (LVAD), where early post-operative changes in sCr correlated with changes in muscle mass as assessed by PMI^24^. Notably, in the same report, sCr-based eGFR showed a transient improvement after LVAD implantation, whereas cysC-based eGFR remained stable. In the early post-HT phase, characterized by muscle wasting and reduced protein intake, sCr may underestimate the degree of kidney dysfunction resulting from surgical insult and acute calcineurin inhibitor nephrotoxicity. Subsequent improvements in the quality and quantity of muscle mass as well as in nutritional status could account for the divergent trajectories of eGFRsCr and eGFRcysC. Factors influencing cysC levels independently from kidney function are also likely to explain variations in eGFRdiff_cysC-sCr_. The association of higher BMI with more positive eGFRdiff_cysC-sCr_ values may reflect the influence of adipose tissue on cysC generation^34^ or the effect of muscle mass on sCr levels. The modest association of prednisone dose with eGFRdiff_cysC-sCr_ in our population is consistent with prior studies demonstrating a steroid-induced, dose-dependent cysC increase not related to kidney function^17,18^.Overall, our findings indicate a large impact of non-GFR determinants of cysC and sCr in the early post-operative period, particularly in light of improved convergence of the estimates at 1-year post-HT.

Establishing an evidence-based approach to assess kidney function pre- and post-HT is critical for current clinical practice. Since the 2018 implementation of the new United Network for Organ Sharing allocation system, patients demonstrate higher acuity at the time of HT, as evidenced by increased rates of mechanical ventilation, intensive care unit stay and temporary mechanical circulatory support, and are more likely to require post-operative kidney replacement therapy^35–37^. The current paradigm for kidney function assessment, which relies heavily on sCr, might have its limitations further accentuated in a patient cohort with a rising incidence of critical illness. Additionally, a new Organ Procurement and Transplantation Network policy restricted simultaneous heart-kidney transplant to patients with an GFR <30 ml/min/1.73 m^2^ and introduced a “safety net” in the presence of persistent kidney dysfunction (eGFR <20 ml/ min/1.73 m^2^, or need for kidney replacement therapy) within the first year following HT^15,38^. Notably, the policy provided no guidance on which marker or equation should be used to calculate eGFR. The significant variations observed in kidney function estimates in the present study may have relevant implications for clinical practice if cysC were to become routinely used. This makes our findings timely and pertinent to the ongoing discussion regarding kidney transplantation eligibility among HT candidates and recipients^6,15,39^.

Valganciclovir dosing recommendations using the Cockcroft-Gault equation are based on the VICTOR trial^40^, a study conducted nearly two decades ago. There are numerous limitations to the Cockcroft-Gault equation with respect to accurately estimating drug clearance. Developed in 1973 using data from 249 men, this equation was not adjusted for body surface area. Also, since the publication of VICTOR trial, sCr assays have been standardized to reference methods. Contemporary sCr-based equations, but not Cockcroft-Gault equation, have been expressed for these reference methods. Given that the assay used in the derivation of Cockcroft-Gault equation yielded higher sCr levels than current methods^41^, its application may lead to higher drug dosage recommendations than originally intended in pharmacokinetic studies. Using cysC, we identified a significant number of HT recipients who were potentially *overdosed* on VGC. Future studies could leverage this pilot data to directly compare VGC dosing using cysC-based equations against other kidney function estimates and evaluate differences in rates of leukopenia and CMV infection.

Our study has several limitations. First, the absence of a gold-standard measure of GFR prevents us from determining which marker offers the most precise kidney function estimate. The single-center design represents another limitation, as our practices and transplantation eligibility criteria might not be representative of other institutions. The number of missing samples at certain time points, particularly pre-HT, raises concerns about potential unmeasured confounders affecting our findings on longitudinal eGFR changes. Nevertheless, our sensitivity analyses on paired samples corroborated the conclusions of the main analysis. We must also acknowledge that the eGFR equations used in this study were initially derived from patients with stable kidney function, limiting their reliability when applied to acutely ill patients. Lastly, although all the findings of the present study are hypothesis-generating, the analysis on leukopenia events is exploratory in nature and should be interpreted with particular caution.

In conclusion, among HT patients, we found significant discrepancies between cysC- and sCr-based kidney function estimates, which could be attributed to changes in body composition and steroid use. These findings may have relevant implications for VGC dosing and other aspects of clinical management. Further research is warranted to define a new standard for kidney function assessment in this patient population.

## AUTHORS’ CONTRIBUTIONS

Conception and Design: AP, DLJ, PCC, MY

Statistical Analysis: AP, RTD, BB

Analysis and Interpretation of the Data: AP, AL, PD, AOS, IM, BB, RTD, KT, GTS, NU, JSL, SAH, SM

Supervision or Mentorship: PCC, MY

## Data Availability

All data produced in the present study are available upon reasonable request to the authors

## ACKNOWLEDGMENTS

None

