## Supplementary Material for "Kidney Function Assessment Using Cystatin C and Serum Creatinine in Heart Transplantation Recipients: Implications for Valganciclovir Dosing"

**Table of contents**

Supplementary Figures – Page 2

Supplementary Tables – Page 5

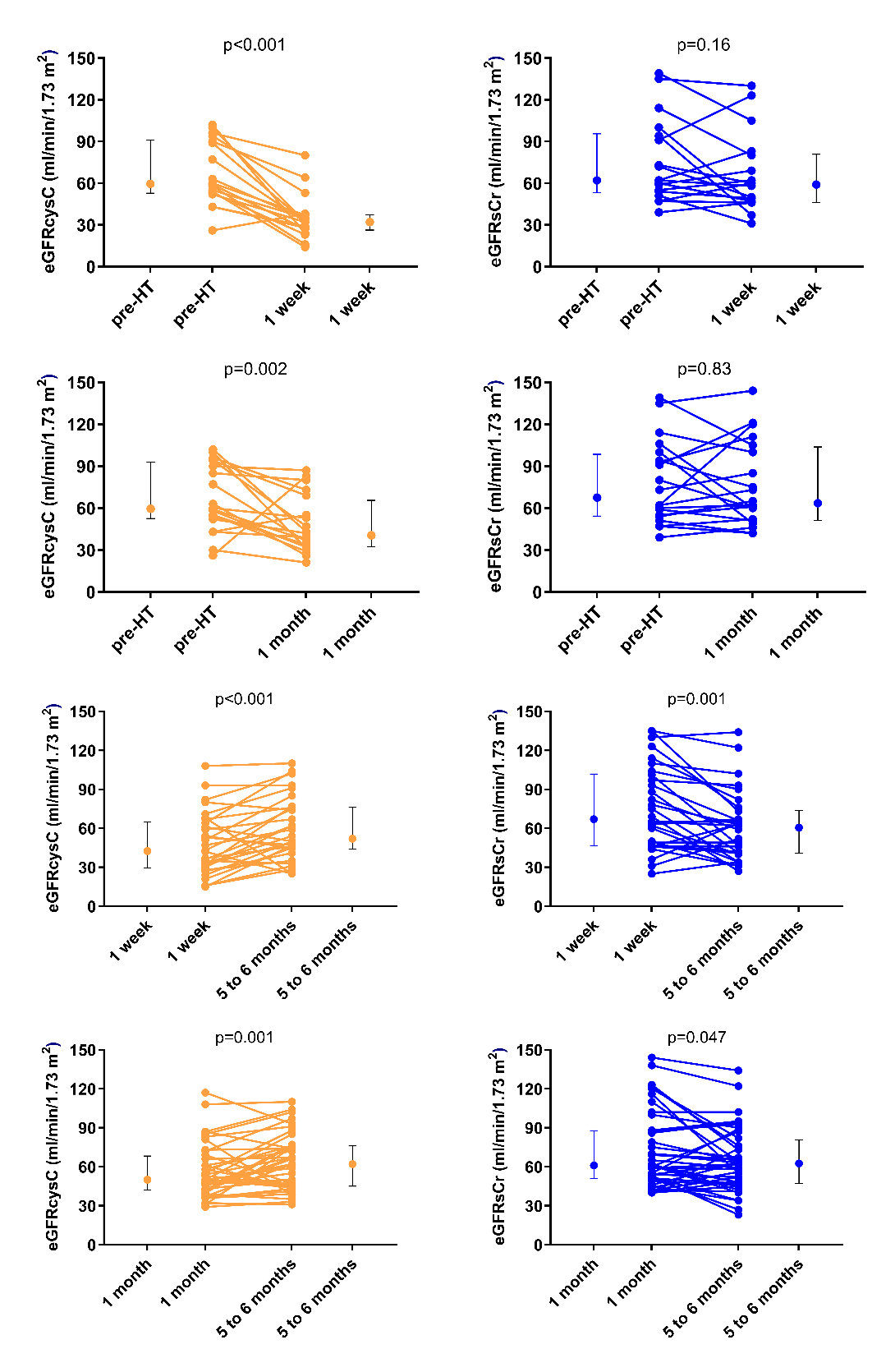

**Figure S1.** Changes in estimated glomerular filtration rate among patients with samples available at both pre-HT and 1-week (or 1-month) post-HT and at both 1-week (or 1-month) and 5 to 6 months post-HT. Bars represent median and interquartile range. cysC indicates cystatin C; eGFR, estimated glomerular filtration rate; sCr, serum creatinine.

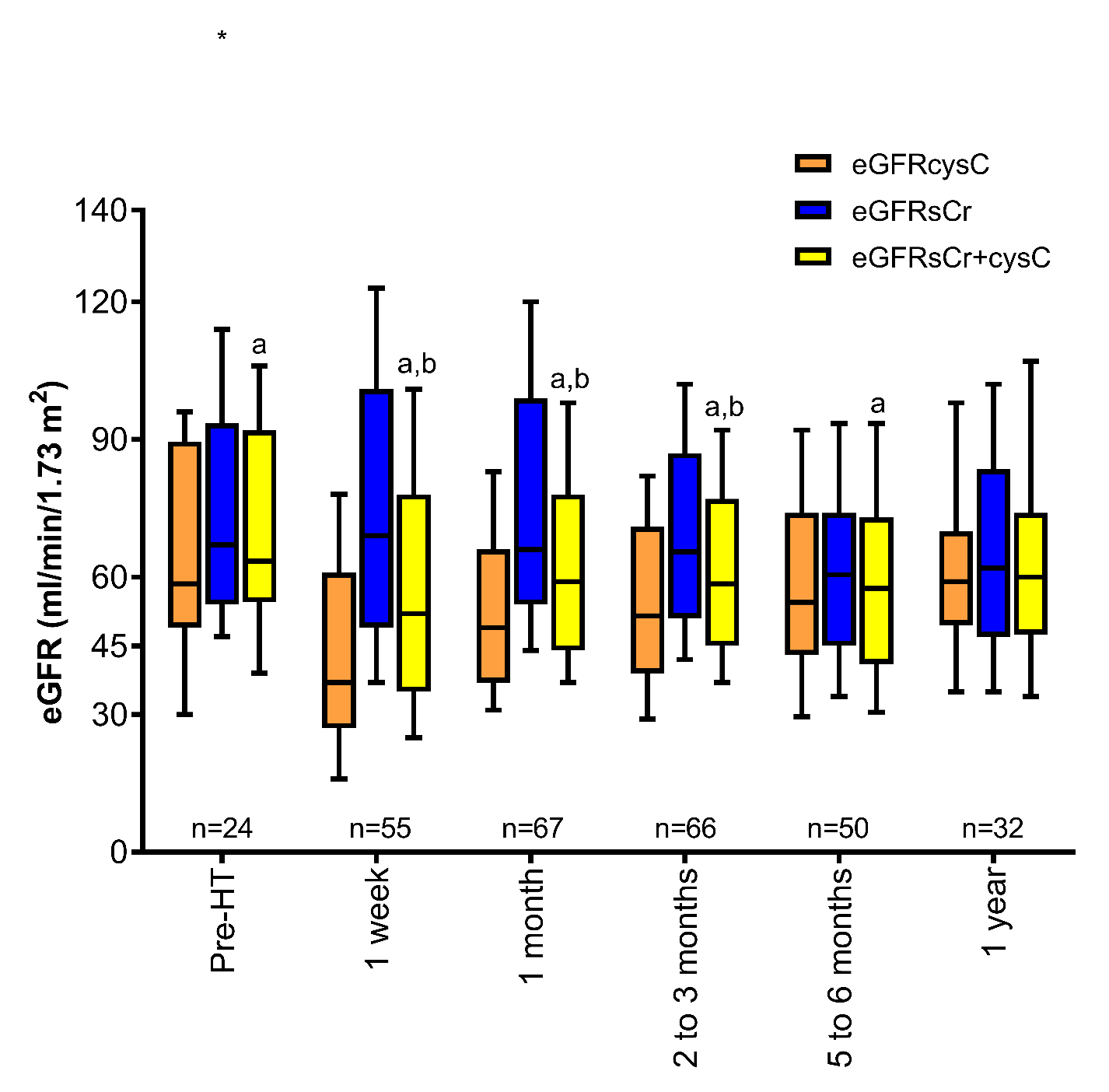

**Supplemental Figure 1.**

**Figure S2.** Kidney function estimates across time points. Boxes and whiskers represent the interquartile range and the 10th–90th percentile range, respectively. Comparisons are shown for the equation combining serum creatinine and cystatin (eGFRsCr+cysC) as follows: a: p<0.05 vs. eGFRcysC; b: p<0.05 vs. eGFRsCr. cysC indicates cystatin C; eGFR, estimated glomerular filtration rate; HT, heart transplant; sCr, serum creatinine.

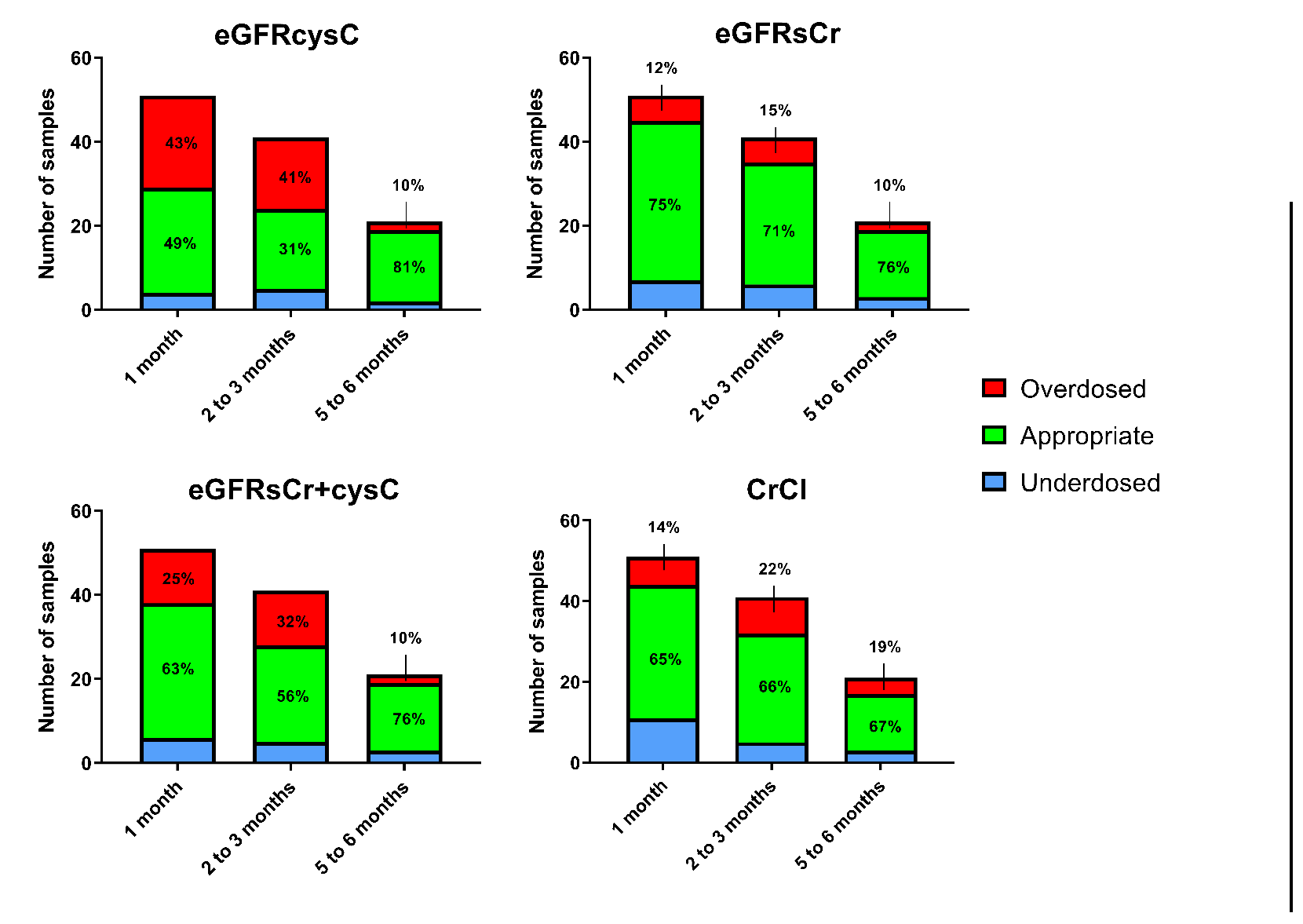

**Figure S3.** Dose adjustments based on estimates of kidney function compared to administered dose stratified by time of sampling. CrCl indicates creatinine clearance; cysC, cystatin C; eGFR, estimated glomerular filtration rate; sCr, serum creatinine.

**Table S1.** Median values of kidney function estimates and of the difference between cystatin C- and serum creatinine-based estimated glomerular filtration rate (eGFRdiffcysC-sCr) at different time points

|  | Pre-HT | Post-HT  1 week | Post-HT  1 month | Post-HT  2 to 3 months | Post-HT  5 to 6 months | Post-HT  1 year |
| --- | --- | --- | --- | --- | --- | --- |
| eGFRcysC | 58.5  b,c,1,2 | 37  b,c,0,1,2,3,4,5 | 49  b,c,0,1 | 51.5  b,c,1 | 54.5  c,1 | 59  1 |
| eGFRsCr | 67  a,4 | 69  a,c,4 | 66  a,c,4,5 | 65.5  a,c,4 | 60.5  0,1,2,3 | 62  2 |
| eGFRdiff_cysC-sCr_ | -10  1,2 | -28  0,1,2,3,4,5 | -22  0,1,4,5 | -15.5  1,4,5 | -2.5  1,2,3 | -2.5  1,2,3 |
| eGFRsCr+cysC | 63.5  a,1,4 | 52  a,b,0,2,3,5 | 59  a,b,1 | 58.5  a,b,1 | 57.5  a,0 | 60  1 |

All values are reported in ml/min/1.73 m^2^. Comparisons are shown as follows: a: p<0.05 vs. eGFRcysC; b: p<0.05 vs. eGFRsCr; c: p<0.05 vs. eGFRsCr+cysC; 0: p<0.05 vs. Pre-HT; 1: p<0.05 vs. 1-week post-HT; 2: p<0.05 vs. 1-month post-HT; 3: p<0.05 vs. 2 to 3 months post-HT 4: p<0.05 vs. 5 to 6 months post-HT; 5: p<0.05 vs. 1-year post-HT. P-values were obtained using signed-rank tests and linear mixed models adjusted for age and sex, as appropriate. No adjustment was made for multiple comparisons.

**Table S2.** Reclassification of estimated glomerular filtration rate (eGFR) categories based on cystatin C at individual time points

| eGFRcysC vs.  eGFRsCr (reference) | Worse eGFR category | Same eGFR category | Better eGFR category |
| --- | --- | --- | --- |
| Pre-HT | 10 (42%) | 12 (50%) | 2 (8%) |
| 1 week | 48 (87%) | 7 (13%) | 0 (0%) |
| 1 month | 48 (72%) | 16 (24%) | 3 (4%) |
| 2 to 3 months | 35 (53%) | 25 (38%) | 6 (9%) |
| 5 to 6 months | 17 (34%) | 23 (46%) | 10 (20%) |
| 1 year | 13 (41%) | 11 (34%) | 8 (25%) |
| Total | 171 (58%) | 94 (32%) | 29 (10%) |

Data are presented as number of samples and percentage of the total samples (%). cysC indicates cystatin C; eGFR, estimated glomerular filtration rate; HT, heart transplant; sCr, serum creatinine.

**Table S3.** Reclassification of estimated glomerular filtration (eGFR) categories based on the equation combining serum creatinine and cystatin at individual time points

| eGFRsCr+cysC vs.  eGFRsCr (reference) | Worse eGFR category | Same eGFR category | Better eGFR category |
| --- | --- | --- | --- |
| Pre-HT | 5 (21%) | 17 (71%) | 2 (8%) |
| 1 week | 39 (71%) | 16 (29%) | 0 (0%) |
| 1 month | 28 (42%) | 36 (54%) | 3 (4%) |
| 2 to 3 months | 22 (33%) | 40 (61%) | 4 (6%) |
| 5 to 6 months | 10 (20%) | 33 (66%) | 7 (14%) |
| 1 year | 7 (22%) | 20 (62%) | 5 (16%) |
| Total | 111 (38%) | 162 (55%) | 21 (7%) |

Data are presented as number of samples and percentage of the total sample (%) at individual time points. cysC indicates cystatin C; eGFR, estimated glomerular filtration rate; HT, heart transplant; sCr, serum creatinine.

**Table S4.** Association of body mass index and prednisone dose with kidney function estimates and the difference between cystatin C- and serum creatinine-based estimated glomerular filtration rate (eGFRdiff_cysC-sCr_)

|  | BMI | | Prednisone dose | |
| --- | --- | --- | --- | --- |
| Model | Estimate | p-value | Estimate | p-value |
| eGFRcysC | -0.162 | 0.21 |  |  |
| eGFRsCr | -0.3843 | <0.001 |  |  |
| eGFRsCr+CysC | -0.2732 | 0.019 |  |  |
| eGFRdiff_cysC-sCr_ | 0.2223 | 0.029 |  |  |
| eGFRcysC |  |  | -0.0758 | 0.14 |
| eGFRsCr |  |  | 0.0794 | 0.079 |
| eGFRsCr+cysC |  |  | -0.0074 | 0.87 |
| eGFRdiff_sCr-cysC_ |  |  | -0.1552 | <0.001 |
| eGFRcysC | -0.2184 | 0.11 | -0.0898 | 0.081 |
| eGFRsCr | -0.4202 | <0.001 | 0.0524 | 0.24 |
| eGFRsCr+cysC | -0.3235 | 0.01 | -0.0282 | 0.55 |
| eGFRdiff_cysC-sCr_ | 0.2018 | 0.053 | -0.1422 | <0.001 |

Estimates reflect changes in log-transformed estimated glomerular filtration rate for each logarithmic increment in body mass index and prednisone dose. All models are adjusted by age, sex, and time point. cysC indicates cystatin C; eGFR, estimated glomerular filtration rate; sCr, serum creatinine.

**Table S5.** Baseline characteristics and drug regimens of study participants stratified by availability of pectoralis muscle measures

|  | Pectoralis muscle measures | |
| --- | --- | --- |
|  | Available  n=21 | Not available  n=59 |
| Demographics and clinical characteristics | | |
| Age, years | 60 [50 to 62] | 56 [47 to 65] |
| Female, n (%) | 6 (29%) | 10 (17%) |
| Black, n (%) | 3 (14%) | 19 (32%) |
| Hypertension, n (%) | 11 (52%) | 22 (37%) |
| Diabetes mellitus, n (%) | 5 (24%) | 13 (22%) |
| Ischemic etiology, n (%) | 8 (38%) | 15 (25%) |
| BMI, kg/m^2^ | 27.4 [23.6 to 30.7] | 26.4 [22.5 to 28.7] |
| LVAD as BTT, n (%) | 57 (71%) | 8 (33%) |
| Baseline laboratory values obtained within 24 hours prior to HT | | |
| Creatinine, mg/dL | 1.20 [0.98 to 1.43] | 1.18 [0.87 to 1.46] |
| eGFR, ml/min/1.73 m^2^ | 63 [54 to 89] | 72 [49 to 95] |
| BUN, mg/dL | 18 [15 to 23] | 18 [15 to 24] |
| WBC, 10^9^/L | 8.0 [7.3 to 8.9] | 7.1 [5.8 to 8.9] |
| During HT admission | | |
| Kidney replacement therapy, n (%) | 4 (19%) | 1 (2%) |
| ECMO for primary graft dysfunction, n (%) | 5 (24%) | 11 (19%) |
| Medications at hospital discharge after HT | | |
| Prednisone, n (%) | 21 (100%) | 58 (98%) |
| Prednisone dose, mg | 25 [20 to 25] | 25 [20 to 25] |
| Tacrolimus, n (%) | 21 (100%) | 57 (97%) |
| Tacrolimus daily dose, mg | 10 [7 to 14] | 6 [10 to 13] |
| Cyclosporine, n (%) | 0 (0%) | 1 (2%) |
| Cyclosporine daily dose, mg | . | 325 |
| Mycophenolate mofetil, n (%) | 19 (91%) | 54 (92%) |
| Mycophenolate mofetil daily dose, mg | 3000 [2000 to 3000] | 3000 [3000 to 3000] |
| Mycophenolate sodium, n (%) | 2 (10%) | 3 (5%) |
| Mycophenolate sodium daily dose, mg | 1260 [1080 to 1440] | 1440 [1420 to 1440] |
| Trimethoprim/sulfamethoxazole, n (%) | 9 (43%) | 36 (61%) |
| Valganciclovir, n (%) | 15 (71%) | 44 (75%) |
| Valganciclovir daily dose, mg | 900 (450 to 900) | 900 (450 to 900) |

Values are presented as median [interquartile range] unless specified. BMI indicates body mass index; BTT, bridge to transplant; BUN, blood urea nitrogen; ECMO, extracorporeal membrane oxygenation; eGFR, estimated glomerular filtration rate; HT, heart transplant; LVAD, left ventricular assist device; WBC, white blood cell.

**Table S6.** Baseline characteristics and drug regimens of study participants stratified by inclusion in the valganciclovir analysis

|  | Included in valganciclovir analysis | |
| --- | --- | --- |
|  | Yes  n=57 | No  n=23 |
| Demographics and clinical characteristics | | |
| Age, years | 56 [47 to 65] | 55 [47 to 65] |
| Female, n (%) | 12 (21%) | 4 (17%) |
| Black, n (%) | 20 (35%) | 2 (9%) |
| Hypertension, n (%) | 25 (44%) | 8 (35%) |
| Diabetes mellitus, n (%) | 12 (21%) | 6 (26%) |
| Ischemic etiology, n (%) | 16 (28%) | 7 (30%) |
| BMI, kg/m^2^ | 27.6 [23.5 to 30.6] | 26.7 [24.9 to 31.2] |
| LVAD as BTT, n (%) | 43 (75%) | 14 (61%) |
| Baseline laboratory values obtained within 24 hours prior to HT | | |
| Creatinine, mg/dL | 1.16 [0.85 to 1.43] | 1.22 [1.05 to 1.46] |
| eGFR, ml/min/1.73 m^2^ | 73 [50 to 95] | 65 [54 to 77] |
| BUN, mg/dL | 17 [15 to 23] | 21 [17 to 25] |
| WBC, 10^9^/L | 7.8 [5.8 to 8.9] | 7.6 [6.9 to 8.6] |
| During HT admission | | |
| Kidney replacement therapy, n (%) | 3 (5%) | 2 (9%) |
| ECMO for primary graft dysfunction, n (%) | 7 (30%) | 9 (16%) |
| Medications at hospital discharge after HT | | |
| Prednisone, n (%) | 57 (100%) | 22 (96%) |
| Prednisone dose, mg | 25 [20 to 25] | 25 [20 to 25] |
| Tacrolimus, n (%) | 55 (96%) | 23 (100%) |
| Tacrolimus daily dose, mg | 10.5 [7 to 14] | 7.5 [5 to 12.5] |
| Cyclosporine, n (%) | 1 (2%) | 0 (0%) |
| Cyclosporine daily dose, mg | 325 | . |
| Mycophenolate mofetil, n (%) | 54 (95%) | 19 (83%) |
| Mycophenolate mofetil daily dose, mg | 3000 [3000 to 3000] | 3000 [2500 to 3000] |
| Mycophenolate sodium, n (%) | 3 (5%) | 2 (9%) |
| Mycophenolate sodium daily dose, mg | 1420 [1080 to 1440] | 1440 [1440 to 1440] |
| Trimethoprim/sulfamethoxazole, n (%) | 30 (53%) | 15 (65%) |
| Valganciclovir, n (%) | 56 (98%) | 3 (13%) |
| Valganciclovir daily dose, mg | 900 (450 to 900) | 900 (950 to 900) |

Values are presented as median [interquartile range] unless specified. BMI indicates body mass index; BTT, bridge to transplant; BUN, blood urea nitrogen; ECMO, extracorporeal membrane oxygenation; eGFR, estimated glomerular filtration rate; HT, heart transplant; LVAD, left ventricular assist device; WBC, white blood cell.
